# The Influence of Pregnancy on PrEP uptake and Adherence Amongst HIV-Negative High-Risk Young Women in Kampala, Uganda: A Qualitative Assessment

**DOI:** 10.1101/2022.04.12.22273506

**Authors:** Shivali Joshi, Catherine Namuddu, Francis Xavier Kasujja, Miriam Mirembe, Jaco Homsy, Janet Seeley, Rachel King

## Abstract

**Background:** Pregnant young women who engage in high-risk sexual activity are at elevated biological and social risk for HIV acquisition. PrEP serves as an effective means of HIV prevention, including during pregnancy. This study aimed to explore attitudes, experiences and challenges with PrEP to understand what motivates or limits PrEP uptake and adherence during pregnancy among this population of young women.

**Methods:** Semi-structured interviews were conducted with 23 participants, recruited from the Prevention on PrEP (POPPi) study in the Good Health for Women Project clinic in Kampala, Uganda. POPPi’s inclusion criteria comprised of HIV-uninfected women, aged 15-24, who engaged in high-risk sexual activity. Interviews focused on experience with PrEP and pregnancy. Data were analyzed utilizing a framework analysis approach.

**Findings:** Key themes were comprised of participant barriers to and facilitators of PrEP uptake and adherence. Reasons for PrEP initiation included desire for autonomy and agency, mistrust of partners, and social support. Participants expressed challenges with initiating or sustaining their use of PrEP, including PrEP access and perceived or felt stigma. During pregnancy, participants’ primary motivators for altering PrEP use were either understanding of PrEP safety for their baby or changes in perceptions of their HIV risk. Many of these factors were similar across participants who had experience with pregnancy and those who did not.

**Interpretation:** This study highlights the importance of addressing barriers to and facilitators of PrEP adherence, especially during pregnancy where risk is elevated, with a multi-level approach. Community-oriented education, stigma reduction activities alongside access to PrEP, can serve as means for adherence. The development of robust PrEP adherence support guidelines regarding PrEP use during pregnancy among high-risk women, and strategies for their implementation, are of utmost importance for the control of HIV in key populations and the elimination of mother-to-child transmission of HIV.

## INTRODUCTION

There have been substantial advances in HIV testing and treatment globally. However, despite these gains, there were 1.7 million new HIV infections worldwide in 2019 before the global COVID pandemic, of which almost 60% occurred in sub-Saharan Africa.(2)

The Joint United Nations Programme on HIV/AIDS defines transactional sex as “a non-marital, non-commercial sexual relationship motivated by an implicit assumption that sex will be exchanged for material support or other benefits”. (3, 4) Transactional sex is different from formal sex work as the relationship or exchange between individuals may not be formally established; it is inclusive of relationships with shared emotional intimacy as well as exchanges of money and goods for sex.(2) In the case of formal sex work, sex is explicitly suggested as a commercial commodity. Much social research on HIV has not included women who engage in sex work of *all kinds*, often due to a lack of categorization of transactional sex practices. Thus, data regarding female sex workers (FSWs), a commonly used classification, often underestimates the number of women who engage in similar sexual behaviors.(2) Here we refer to both formal sex workers and women who engage in transactional sex, as young women who are high-risk (YWHR).

In Uganda, women who engage in both transactional and non-transactional sex have one of the highest risks for HIV acquisition. This is the case even in comparison to other high-risk groups including men who have sex with men, injection drug users, and prisoners.(5) HIV prevalence amongst FSWs is close to 33%.(5, 6) FSW in Uganda reside mostly in urban centers as well as some rural regions of the country. Because sex work is illegal in Uganda, they must operate amongst informal sectors of society. (7) FSWs, can be prosecuted and incarcerated even without strong evidence.(7) As a result, they face a number of challenges to well-being relating to criminalization, violence, mobility, stigma, and other abuse.(8) Legal barriers, along with cultural and religious constraints, perpetuate stigma and social isolation. Thus, navigation of reproductive and sexual health resources remains extremely difficult for FSW in Uganda.

While these vulnerabilities are true for high-risk women of all ages, the vulnerability of young women is heightened. Young women new to sex work are at higher risk for contracting HIV and other sexually transmitted infections (STIs).(9) Biologically, younger women possess an increased risk for trauma upon sexual contact compared to older women. There are some possible explanations from Dellar *et al* (2015) such as an immature cervix with the exposure of a higher proportion of genital mucosa with high levels of genital inflammation.(10) Behaviorally, higher incidence of drug use has been associated with YWHR.(9) Socially and culturally, societal pressures are harder to navigate for younger, less powerful members of society. (10) Additionally, due to the illegality of sex work in Uganda, YWHR are forced to work and live in areas of cities that are least equipped with adequate health resources.(9) As a result, HIV prevention services for YWHR are limited, and there is a need for interventions that address the specific sociopolitical and cultural barriers that affect this population.

The Prevention on Pre-exposure Prophylaxis (POPPi), the parent study of this sub-study, has examined YWHR in Uganda amongst their social and sexual networks.(11) POPPi aimed to empower YWHR with effective HIV prevention tools that they were able to control and administer themselves, namely Pre-exposure Prophylaxis (PrEP) and HIV oral fluid self-tests.(11) The study focused on this young population to both intervene early, before women are infected, and provide services to a group that maybe overlooked or have difficulty accessing healthcare.

Little research has been conducted on adherence to PrEP once YWHR become pregnant. Addressing uptake and adherence among such women is of critical concern because many YWHR continue to work during early stages of pregnancy to make money before they are no longer able to work. In addition, pregnancy itself is a vulnerable time for contracting HIV as it causes biological changes, including alterations in levels of progesterone and estrogen and the vaginal environment, which are known risk factors for HIV infection.(12, 13) Additionally, pregnancy modifies innate and adaptive immunity, creating a lowered immune response throughout pregnancy. Contracting HIV during pregnancy also increases risk for maternal-to-child transmission; thus, understanding the determinants of HIV prevention and PrEP adherence during pregnancy is critical for both pregnant YWHR mothers and their children.(13)

The WHO acknowledges concerns regarding PrEP and potential adverse birth outcomes, however strongly recommends Tenofovir-based PrEP for pregnant, high-risk populations.(1) There is consensus that the risks of contracting HIV during pregnancy, birth and breastfeeding far outweigh the largely unknown risks of PrEP use.(14) There are a number of policy and health-systems barriers that affect the way PrEP is used prior to and during pregnancy. These barriers can create uncertainty regarding the safety of PrEP for pregnant women, as this information is often not readily available to the public and/or enforced within health facilities and care protocols.(14) Social level barriers surrounding PrEP stigma and misconceptions affect PrEP acceptance and access, as taking PrEP is often equated to taking anti retroviral medication when already infected with HIV. For populations who are wary of being exposed in terms of occupation or sexuality, this misconception makes it risky to be seen taking the pill.(1) Taking PrEP may also imply that one is actively engaging in sexual activity. As a result, taking PrEP for individuals in partnerships could suggest unfaithfulness and women more often than men may be accused of this.(1) Lastly, personal barriers, including education to assess the risks and benefits of taking PrEP and adhering to it while pregnant and breastfeeding, must be considered.(1) Thus, evaluation of the multi-level barriers of using PrEP during pregnancy and the postpartum period, is crucial to understanding attitudes towards PrEP and PrEP adherence among a high risk population. As part of the POPPi study, the present sub-study explored the way these barriers impact YWHR in Uganda, along with general attitudes of YWHR towards PrEP in hopes to understand what motivates PrEP adherence, or lack of, during pregnancy.

## MATERIALS AND METHODS

The goal of the parent study, (POPPi) was to develop and assess an intervention to enhance initiation and adherence to PrEP among HIV negative Young Women at High Risk which began recruitment in February 2019 and completed a 12-month follow-up in 2020.

In 2017, the study team mapped out hot spots and popular meeting points frequented by young women and high-risk youth in urban Kampala, the capital city of Uganda (population about 2.5 million). Phase 1 of POPPi consisted of conducting focus group discussions (FGDs) and interviewing participants to both understand the use of HIV prevention services and identify the overall feasibility and acceptability of the proposed behavioral intervention to improve uptake and adherence to PrEP. Focus groups were held with community members, YWHR, key informants, and peer educators, and interviews were specifically conducted with key-informants at the study clinic. Phase 2 of the study was a pilot randomized controlled trial (RCT) comparing POPPi intervention outcomes to the study clinic’s standard of care. This intervention consisted of group sessions on health empowerment, PrEP literacy, and social empowerment, along with individual counseling sessions on topics like HIV and STI transmission, and family planning. Phase 2 included a 12-month follow-up, to evaluate PrEP uptake and adherence using self-report and a biomarker of PrEP drug levels in hair.

### Sub-Study Design

This sub-study employed a qualitative methodology given the nature of the research question, which focused on informing elements such as health-seeking behavior, decision-making, and motivations that were best captured through a flexible form of data collection, specifically group discussions and semi-structured interviews aimed to gather views on focused topics, while remaining open to accommodate new and emerging subjects and themes in a private and confidential manner.

The recruitment process and data collection started in February 2020, data collection continued through June 2020.

### Study Participant Selection and Recruitment

The study population of interest was the same as the parent study (POPPi): young women who engage in high-risk sexual activity as defined by the behavior that put them at risk of contracting HIV, other STIs, or getting pregnant, a definition that is used within the study clinic where other studies on this population were implemented. The clinic recruitment questions included:

> *Have you been on a fishing landing site in the past 3 months? Have you had an STI in the last 3 months? How many times do you have sex with different partners a day? How many irregular and regular sexual partners do you currently have? How many times do you use condoms with those different partners?*

Women were recruited from bars and lodges in Kampala, where commercial sex was available, and subsequently screened for HIV, hepatitis B, and creatinine levels. They were then enrolled in POPPi at the study clinic. For the purposes of this sub-study, potential participants were identified by the POPPi research team and contacted by phone, based on their knowledge and rapport with POPPi participants and their ability to assess eligibility.

Participant eligibility mirrored POPPi’s inclusion criteria, as follows: women aged 15-24 years currently enrolled in POPPi in Kampala, Uganda, testing HIV-antibody negative at enrolment in this study, having demonstrated understanding of POPPi study guidelines and scope, and providing oral and written consent. Eligible women were recruited into one of four categories to acquire relevant themes for data analysis: (1) PrEP-naive, (2) having started and stopped PrEP, (3) having started, stopped, and reinitiated PrEP, and (4) stable on PrEP. Of these, women who were currently pregnant or had previously been pregnant while enrolled in the study, were contacted first. Nulliparous women were contacted to supplement as needed to fill the four categories of PrEP. No specific pregnancy duration or timeframe was used given the young age of participants implying any past pregnancy likely occurred fairly recently.

### Data collection

Interviews began in February 2020 and were conducted by an experienced social science interviewer, in Luganda either at the study clinic or at participants’ homes. Participants were compensated 20,000 Uganda shillings (about US $6) for their time. Participants were identified solely by their study IDs; no identifiable data were recorded. De-identified data were securely stored on an encrypted device and audio recordings were deleted after transcription was completed.

A total of 23 interviews were conducted and analyzed reached thematic saturation.(15) Interviews followed a semi-structured guide which contained an introduction to the study, asked participants for a brief background including their sexual and reproductive history and characteristics, and categorized specific questions and probes by modes of inquiry. The two major modes of inquiry, experience with PrEP and PrEP and pregnancy, asked participants about motivations to take PrEP, knowledge on the prevalence of PrEP use in the community, personal PrEP use and adherence during pregnancy, and types of care and support available while pregnant.

### Data analysis

Audio recordings of the interviews were uploaded onto a secure device, first transcribed in Luganda, and subsequently translated to English. A framework analysis approach was used for data analysis (Figure 2).(16) The analytical framework in this approach allowed the researchers to map and categorize data as well as find associations and ultimately derive conclusions.(17) Transcripts were uploaded to NVivo, a qualitative data analysis software, where they were coded. A codebook was developed, defining each of the codes, both broader, thematic codes, and sub-codes that fell under the themes.(16) The codes were then re-applied to the transcripts in NVivo, with heightened scrutiny and attention. A framework analysis matrix was developed in Microsoft Excel using major themes as categories and summarizing each transcript by category. These summaries contained quotes, observations, and overall content summarization. Once the data was in the framework analysis matrix, it was interpreted and used to derive associations, interpretations and conclusions.(16) The preliminary findings were summarized and presented both to study staff members and a selected group of study participants.

#### Ethical considerations

All participants provided written informed consent and understanding was assessed. We included some minors in this study (seven in total). As many young sex workers are actually orphans, in agreement with the Uganda National Council of Science and Technology, all minors who fit with the definition of ‘emancipated minors’ were eligible to consent to research without their parents also consenting.

## RESULTS

### Participants

Almost half (48%) of the participants fell between the ages of 19-21 years, (Table 1). In terms of educational background, Uganda’s education system includes seven years of primary level education and six years of secondary level, followed by tertiary or university education.(18) Most participants had left school, due to a lack of financial capability to continue education, either during Primary or Secondary school. The majority of participants declared themselves Roman Catholics or Muslims (30% and 22%).

**Table 1.** Participants’ demographic characteristics, Kampala, Uganda

| *Participant Characteristics (n=23)* |  |  |
| --- | --- | --- |
| *Characteristics* | *Frequency (n)* | *Percent (%)* |
| **Age** |  |  |
| 15-18 | 7 | 30% |
| 19-21 | 11 | 48% |
| 22-24 | 5 | 22% |
| **Highest formal education** |  |  |
| Started Primary | 7 | 30% |
| Completed Primary | 4 | 17% |
| Started Secondary | 11 | 48% |
| Completed Secondary | 1 | 4% |
| **Religion** |  |  |
| Born Again Christian | 4 | 17% |
| Catholic | 7 | 30% |
| *Church of Uganda-Anglican* | 3 | 13% |
| *Muslim* | 5 | 22% |
| *Adventist* | 1 | 4% |
| *Hindu* | 1 | 4% |
| *Unknown* | 2 | 9% |

With regards to sexual and reproductive history and characteristics, most participants had either one or no children. The majority had between one and nine different sexual partners within a one-month period (Table 2). When asked about condom use with spouses or partners they lived with and with paying customers within the past month, most participants said they *never* used male condoms with partners, while with paying customers the majority of participants *sometimes* used male condoms (Table 2). The two most common reasons for not using male condoms during sexual interactions with either partners or paying customers were: 1) sexual partners preferred sex without condoms, and 2) there were no condoms available. Most participants reported not having used addictive substances in their lifetime, however, for those who did, the substances consumed were shisha (tobacco), mayirungi (khat), and cocaine (Table 2).

**Table 2.** Participants’ sexual and reproductive health characteristics, Kampala, Uganda

| *Participant Sexual and Reproductive Characteristics (n=23)* |  |  |
| --- | --- | --- |
| *Characteristics* | *Frequency (n)* | *Percent (%)* |
| ***Number of children*** |  |  |
| 0 | 12 | 52% |
| 1-2 | 10 | 43% |
| 3-4 | 1 | 4% |
| ***Number of total partners (in the past month)*** |  |  |
| 1-9 partners | 16 | 70% |
| 10-19 partners | 2 | 9% |
| 20-49 partners | - | - |
| Over 50 partners | 2 | 9% |
| No data | 3 | 13% |
| **Condom Use**<br>**(in the past month)** | *With partners \| With paying*<br>*customers* | *With partners \| With paying*<br>*customers* |
| Never | 8 \| 5 | 35% \| 22% |
| Sometimes | 5 \| 8 | 22% \| 35% |
| Most of the time | 1 \| 4 | 4% \| 17% |
| Always | 3 \| 2 | 13% \| 9% |
| **Drug Use (Ever)** |  |  |
| Yes | 7 | 30% |
| No | 15 | 65% |
| No data | 1 | 4% |

Participants were categorized based on PrEP status and parity, as seen in Figure 1. In the text we have identified participants based on age groups, under 20 and 20-24 years old as well as PrEP status. Most participants reported they were *stable on PrEP* and *not currently pregnant*. Many participants had a pregnancy during their lifetimes but, because they did not get pregnant during the course of the POPPi study, were excluded from the *previously pregnant* category. We determined five major themes around barriers to, and facilitators of adherence to PrEP: 1) agency and trust, 2) safety, 3) HIV and sex work-related stigma, 4) social support and societal perceptions, and 5) access to PrEP and PrEP information.

### Agency and trust

Participants’ decisions to initiate PrEP and stay consistent on PrEP were influenced by their desires for autonomy over their sexual health and livelihood. One participant under 20 years old, who was stable on PrEP and pregnant at the time of the interview, expressed her preference for PrEP over other forms of HIV prevention, like condoms:

> *Because with PrEP, it’s you to control it, if you don’t swallow it, it’s you to get infected but for a condom, if it’s to burst, [you have no control over it]*.*”*

She described a sentiment that multiple participants shared, that PrEP enabled a choice to protect one’s life. Thus, upon learning about PrEP, many were motivated to try it.

Many participants also expressed that mistrust in partners influenced their desire to take PrEP. Participants who did not trust their partner’s behavior, in terms of having multiple sexual partners, found PrEP to be a way to ensure their own health, while a few who were consistently with a partner they trusted, did not feel the need to take PrEP. One participant shared a conversation she had with her partner.

“*I explained to him the reason I was swallowing these pills [PrEP], but if he is serious that we are going to be together at home and he is not going to have partners outside our relationship I can stop them because even the basawo [doctors] told me that if a time comes and I get a serious partner and we are going to stay together it is your right to stop swallowing the pills”* (under 20 years old, stable on PrEP, previously pregnant).

This trust, or lack thereof, also extended to partners’ use of condoms. All participants reported experience having tried to use condoms regularly. Participants who were able to ask customers and regular partners to use condoms were less inclined to stay on PrEP. Those who were unable to ensure condom use during sexual interactions were more motivated to take PrEP. Additionally, a few participants also expressed trusting their partners enough not to use condoms. If this was the case, they were still able to use PrEP to maintain continuous protection. As expressed by one participant:

> *“There is only one guy I don’t use a condom with and that’s because when we first met, he told me we go to hospital and test together which we did, and we were both HIV negative. But I still take PrEP whether I’m meeting with this guy or with any other customer”* (under 20 years old, stable on PrEP, previously pregnant).

PrEP allowed women choices within their occupation as well as opportunities for future occupations. Some participants were motivated to take PrEP to stay healthy in the present, so that future work opportunities and pathways of life would remain open. When describing her conversations with other women, one participant shared,

> *“I tell them that we have to adhere to PrEP because we don’t know what’s ahead of us according to the kind of work we do. Let’s take our drugs because it’s for our own good, you never know in future we might decide to quit this work but it is good to quit when we are still negative and [able to] do other things”* (20-24 years old, stable on PrEP, not pregnant).

### Safety

Issues of safety related to personal security, PrEP safety, and occupational safety were the most commonly discussed reasons for initiating or stopping PrEP. Participants expressed PrEP’s ability to provide personal protection even when customers, partners, and society at large could not. In situations where condoms were not an option (customer tore them, refused to wear them, condoms were expired/burst, or not available), participants were still able to keep themselves physically safe.

Participants talked about the inability of society to keep YWHR and the sex worker community safe. If participants were in a situation where force was used, or a customer refused to wear a condom, there was no one to turn to. Thus, it was their own responsibility to ensure their personal safety. Two participants shared these experiences,

> *“At times you link up with a customer and you agree with him that you are going to use a condom and on reaching inside the room he insists he wants live [condomless] sex; when you make an alarm the outsiders laugh”* (20-24 years old, stable on PrEP, previously pregnant).
>
> *“Me, I want condoms but what if he refuses and uses force on you?*…*Remember you’re at his place you have nowhere to report him. You also fear to say you were raped. Me, I fear”* (under 20 years old, stable on PrEP, not pregnant).

The safety or lack of safety of PrEP was a large determinant as to whether participants sought out PrEP and continued to take the drugs. Most participants were aware that PrEP was safe for pregnant women and their unborn babies; however, some participants believed that PrEP would harm the baby. This was the most common reason for not initiating PrEP amongst participants who had experience with pregnancy.

> *“When I was pregnant with this child I was told to take PrEP which I somehow feared but then I accepted to take it, reaching home I again feared and decided to take the pills back to the clinic and told the counselors I would take the pills after pregnancy because I feared that the pills might affect my unborn baby”* (20-24 years old, started, stopped, & re-initiated PrEP, previously pregnant).

One participant feared that taking PrEP during pregnancy would interfere with her high blood pressure, so she stopped. Another was not comfortable with the PrEP-related side effects she experienced during her pregnancy; she also took a break from the drugs. All participants who were on PrEP mentioned side effects as part of their experience with the medication, though in most cases, these effects did not deter participants from initiating or persisting on PrEP. Those who considered PrEP safe and trusted PrEP either had been tested after having unprotected sex and were still HIV negative or had heard from friends that the drug was effective. Another factor that led many participants to stop PrEP use was the safety of high-risk work while pregnant. Though some women who became pregnant continued to work for the first three to five months of pregnancy, there were many who stopped working due to the perceived and real danger of multiple sexual partners while pregnant. Because these women were not working, they stopped taking PrEP.

> *“We get customers who beat us up and kick us, I realized I would get complications and besides they say it’s not good to sleep with different men while pregnant. The child may get complications”* (20-24 years old, never started PrEP, currently pregnant).

### HIV and sex work-related stigma

All 23 participants discussed their experiences with PrEP-related stigma. Because PrEP pills resemble HIV antiretrovirals (ARVs), participants shared that society understood that those who take PrEP are HIV positive, as most did not know the difference between PrEP and ARVs. Thus, the high-risk community on PrEP is stigmatized similarly to individuals who are HIV positive.

> *“I cannot talk to anyone about it [PrEP] because people have different views about the drug. Someone can show you they have understood and accepted yet in actual sense they have not bought the idea and even talk about you to other people telling them you are HIV positive”* (under 20 years old, stable on PrEP, not pregnant).

Most participants were unable to share that they were on PrEP with friends, family, and partners. In many cases, this did not deter participants from initiating PrEP; however, it made it challenging to stay consistent with PrEP. Some participants would take their pills in secret and lie to family and partners about the medication. One participant was going to stay with friends for a while and intentionally left her PrEP at home for fear of her friends finding the PrEP container.

> *“I thought they will think I am HIV infected because if someone knows you’re infected they can’t love you the same way as before”* (under 20 years old, stable on PrEP, not pregnant).

Stigma also played a role in dictating at what point participants had to stop working. A few participants spoke about personal or friends’ experiences working through the first few months of pregnancy but not being able to attract customers once the pregnancy was showing due to stigma, and thus not initiating or persisting taking PrEP because not sexually active.

> *“Once pregnant, we get off the streets*… *it’s because no one can buy a pregnant woman when he is seeing [another] one who is not [pregnant]”* (20-24 years old, stable on PrEP, not pregnant).

Participants spoke of instances regarding stigma in healthcare settings. Many participants went to government hospitals to access free antenatal care. Some descriptions of stigma related to sex work did not emerge as a major indicator of starting or stopping PrEP; however, a participant described the way in which this stigma could lead to difficulties with obtaining PrEP when a high-risk woman was pregnant.

> *“After getting pregnant those girls don’t want to go back where they were accessing PrEP from…There is a neighbor of mine who used to get her medicine from Mulago but when she became pregnant she did not go back”* (under 20 years old, stable on PrEP, previously pregnant).

### Social support and societal perceptions

Participants described the role of social networks and societal perceptions in encouraging initiation into sex work, motivating PrEP use, and influencing community health and safety. Often, participants were recruited into sex work by their friends who noticed that participants were unhappy in their current occupations or without income and saw sex work as an economic opportunity. Thus, many participants followed their friends into sex work, often living together or with groups of women who all worked as sex workers together.

Information on PrEP and HIV prevention traveled through the high-risk community from individual to individual, influencing participant opinions and choices. These perceptions influenced participant behavior as some participants hid PrEP from their friends while others encouraged their fellow sex workers to join studies where they could access PrEP and care. Most participants were told about the POPPi study by other friends and continued to recruit friends throughout their time in the study. An under 20-year-old study participant, stable on PrEP and previously pregnant, described her experience with educating a friend about PrEP.

> *“A friend of mine was so worried that men will infect her with HIV because they don’t want to use condoms and she wanted to know where she could get PrEP, I showed her what the pills look like and told her to search around for them*.*”*

Although participant behavior regarding PrEP was influenced by societal perceptions, social networks did not seem to determine whether participants stopped or stayed consistent on PrEP.

Support for high-risk women, outside of smaller social networks, especially for pregnant women, was unknown to the participants. All participants stated they did not know of any organizations, individuals/communities, or systems for support for pregnant sex workers and high-risk women, aside from those offered at the study clinic. One participant, when asked how she supported herself while pregnant shared,

> *“Sometimes the father of this child would give me financial support. I would be patient and wait in case I used it up. I used to plan, stock food and used the rest to shop for the baby. Feeding was hard, I used not to eat food”* (20-24 years old, never started PrEP, previously pregnant).

### Access to PrEP and PrEP information

As a part of the study clinic and the POPPi study, participants were given access to PrEP according to Uganda’s Ministry of Health guidelines. This allowed participants education on how to use PrEP, when to take it, and how to integrate it into one’s lifestyle. However, amongst larger communities of women engaging in sex work, participants stated that many did not know about the existence of PrEP.

> *“It’s not easy if the hospital has not informed you, however for us we got a chance that we have a clinic which informs us*…*When you look at our government have you ever heard it announce pills preventing HIV…All they do is to give out condoms and that’s why our clinic is a better option, it sensitizes you about new things” (*20-24-year-old, stable on PrEP, previously pregnant)

Along with lack of access to information on PrEP, another participant expressed that it is difficult for women to find PrEP in health facilities and physically access it.

> *“If PrEP is medicine for HIV/AIDs prevention, how come for us, in clinics and pharmacies, we don’t find it*.*” Often, women are unable to travel to go get the medication even if it is provided at government hospitals*. (20-24-year-old, stable on PrEP, currently pregnant)

One participant brought up the potential expense associated with the drug when asked if pregnant women have access to PrEP. While access to PrEP was not a barrier for participants in this study, they communicated that access to PrEP outside of the study would prevent women they knew from taking the medication.

## DISCUSSION

This qualitative study, nested within the POPPi study, aimed to understand major barriers to, and facilitators of PrEP uptake and adherence amongst young, pregnant and non-pregnant, high-risk women in Kampala, Uganda. Most common reasons for PrEP initiation included desire for autonomy and agency, trust and lack of trust of partners, social support, stigma, and societal perceptions of PrEP. Participants expressed a few challenges with initiating or staying consistently on PrEP, including PrEP access and stigma. During pregnancy specifically, participants’ primary motivators for altering PrEP use were either understanding and misunderstanding PrEP safety or changes in beliefs regarding their HIV risk. These findings were organized within the ‘Barriers to PrEP use’ framework adapted from Davies and Heffron, to demonstrate the interrelatedness of PrEP barriers and facilitators and understand them within the personal, societal health systems, and larger policy level actors (Figure 2).(1)

### Personal Level

At the personal level, participant motivations for and challenges with taking PrEP were largely dependent on the way PrEP made individuals feel. For many, the medication provided a feeling of personal protection and control over one’s body and health. Participants were less reliant on partners and customers, and their use of condoms, to ensure their protection from HIV.

PrEP also enabled participants to foresee a future in which they continued to be healthy enough to work and transition into opportunities other than sex work. Previous studies have described low PrEP uptake amongst young, mobile populations due to difficulties with adherence and low perception of risk.(19) However, many participants in POPPi were aware of their vulnerability and high-risk circumstance, and thus, were eager to take PrEP. This may have, in part, been due to this study population’s exposure to HIV education, as lowered risk perception has been seen in populations with less robust HIV education.(20)

Participants who faced personal difficulties with staying on PrEP attributed these to the drug’s side effects. Though these side effects were not unexpected for most and did not deter participants from initiating PrEP, they created anxieties around taking the drugs along with disruptions in their schedules, causing their PrEP adherence to fluctuate. Many participants worked extremely late nights and early mornings, and thus, having to remember to take a pill daily at the same time proved difficult. Both experiences with side effects and challenges with PrEP logistics make a case for longer-acting PrEP formulations and adequate time to adjust to side effects.(20) Participants expressed anticipation of a form of PrEP that could be taken monthly, or every six months. The recent successful performance of injectable PrEP could drastically eliminate many of these barriers for high-risk communities.(21)

A woman’s risk of HIV acquisition doubles during pregnancy and breastfeeding.(22) Though this heightened risk was not a driver of changes in participant engagement in risky behaviors, participants’ understanding of their HIV risk changed throughout pregnancy. Because some participants were unable or unwilling to work throughout pregnancy, due to fear of harm directed at themselves and their child, many stopped taking PrEP. Others worked through the first three to five months of their pregnancies and would take PrEP while sexually active; these participants stopped PrEP when they stopped working. Either way, most were able to accurately identify their risk periods and adjusted their PrEP use accordingly. Understanding risk and perceptions of risk has been known to aid in PrEP consistency and uptake.(23) This feature, demonstrated by the participants, is likely also attributed to robust HIV education through the POPPi study.

### Societal Level

Social perceptions of PrEP were crucial in participants’ understanding of the medication and the way in which participants could take PrEP comfortably. Participants often learned about PrEP from their social networks and were either encouraged or discouraged to initiate the prophylaxis by peers. Social and peer support appeared to be critical to the process of accessing and adhering to PrEP, confirming what has been found in a number of vulnerable communities.(24, 25)

Participants were challenged by both HIV and sex work-related stigma that pervaded society. Most participants said they hid their PrEP use from family, friends, and partners, for fear of being mistaken as an HIV-infected person taking ARV, thus making it difficult to stay consistent on the medication. Nevertheless, participants found creative ways to navigate stigma out of necessity and it did not seem to serve as a barrier for PrEP initiation or continuation.

Partner “buy-in” and trust were other important factors in shaping participants’ PrEP use. Previous studies explored the role of partner influence, often as a barrier to taking PrEP, a theme that emerged in this study as well.(26) In this study, partners directly influenced participants perceptions of the drug itself, as when partners’ misunderstandings or misperceptions of PrEP or sexual history and patterns either hindered or reinforced women participants’ attitudes and practices with PrEP.

### Health Systems Level

Although study participants were uniquely positioned to understand, access and receive care and support about PrEP, they spoke of the health system barriers that impacted their peers’ and communities’ ability to safely access PrEP and the lack of provision, information and support provided by the government and large hospitals regarding PrEP. Previous studies have demonstrated that physical access to PrEP is correlated to PrEP uptake.(26) Though this did not affect participants in this study, physical access is likely a challenge for other members of the YWHR community.

Pregnant or previously pregnant participants also expressed that pregnant YWHR face healthcare stigma in Uganda and a lack of known support systems for pregnant, high-risk women. Most could not name an organization or community space other than the study clinic that provided care for pregnant YWHR. Pregnancy rates are generally very high in Uganda and extremely high amongst the high-risk community in high-HIV burden regions, thus making the lack of support system and structures for this especially vulnerable population a critical gap for

the overall health of young women and the elimination of mother-to-child transmission of HIV in Uganda.(27)

### Policy Level

Pregnant or previously pregnant women in the study faced many of the same challenges as those who did not have pregnancy experience. However, a primary motivator of pregnant participants for declining, stopping, or waiting to initiate PrEP until after pregnancy, was the question of safety of PrEP during pregnancy. The World Health Organization (WHO) recommends offering PrEP to pregnant and breastfeeding women who are at a high risk for HIV acquisition.(22) Though much of PrEP-related literature does not report any adverse effects of PrEP on mother or child throughout pregnancy, this is a highly under-researched area.(27, 28) Due to the prioritization of fetal health, often times over maternal health, clinical research has historically excluded pregnant women from trials relating to PrEP safety. As a result, though many international policies do not exclude pregnant women from the population eligible for PrEP use, safety is still a large concern and the majority of studies call for more research. Uganda’s policy suggests PrEP for the HIV-negative partner in an HIV discordant couple and does not contraindicate PrEP during pregnancy or breastfeeding.(1) Most participants in this study who did not wish to initiate PrEP while pregnant or who stopped PrEP over the course of their pregnancy were worried about potential effects on their babies. Many stated they were taught at some point in their lifetimes that taking drugs while pregnant was not safe for the fetus. This fear is extremely important to consider as pregnancy and breastfeeding are critical times for HIV acquisition.(22) Most participants who were educated about the safety of taking PrEP while pregnant, when asked, recommended that high-risk women stay on PrEP throughout pregnancy. Thus, both additional research and educational interventions are necessary to appropriately address fears surrounding PrEP’s fetal effects.

Recommendations for improvements in PrEP adherence can also be conceptualized within the multi-level barrier and facilitator framework. At the personal level, educational interventions could help YWHR navigate changing life circumstances with regards to partners and pregnancy.(23) At the social level, targeting sexual and social networks is extremely important for interventions, along with education that targets YWHR networks, especially their male partners who are socially positioned not only to alter their own behaviors with regards to PrEP but also to encourage PrEP use amongst their sexual partners.(29) Integrating wider social networks into HIV prevention and PrEP education could aid to reduce stigma.(20) At the health system level, access is of utmost concern. Ensuring affordable and widespread PrEP access at hospitals and clinics in Uganda, followed by community-oriented information and education on PrEP is likely to improve PrEP awareness. Alleviation of barriers such as distance or lack of transportation for YWHR to obtain PrEP should be part of this effort. Communities must be offered PrEP in clinical spaces with educational tools available and robust advocacy. Moreover, national PrEP clinical guidelines with emphasis on PrEP safety during pregnancy are essential. Equally important is to address the grave consequences that stigmatizing and neglecting pregnant YWHR carry in terms of these women’s and their babies’ health.(28) Lastly, though participants in this study did not speak of the criminalization of sex work in Uganda, this has profound implications for PrEP access and use at the policy level and for the eventual elimination of HIV in society.

This study contributes to important insights on PrEP adherence within specific high-risk communities in a sub-Saharan African social and cultural context. It is novel in that it includes the perspectives of pregnant women among a most vulnerable population. Future studies must consider the importance of addressing the perspectives and the needs of the poorest of the poor in the words of late Paul Farmer in order to be able to truly control epidemics.(30) Mobilizing social services, the transactional sex and sex worker communities, and other community based organizing efforts can help fill gaps in research and interventions, blending medical, systemic and structural, and behavioral approaches.

### Limitations

There were a few limitations to this study. As the study included some non-pregnant women, perspectives offered by these participants, especially on questions of pregnancy, may not have been as representative of the pregnant high-risk population. The study also relied on self-report to understand participant’s PrEP use. Thus, participants account of their PrEP use or willingness to disclose PrEP use may have resulted in some inaccurate information. The study provides for a biomarker, analysis of drug levels in hair, that will compare with self report. Another limitation is that the interviews were primarily analyzed by an English-speaking investigator, thus the process of data analysis in English may have resulted in the possibility of misinterpretation of words or ideas, as some participant perspectives may have been lost in translation.

## CONCLUSION

This study highlights the importance of addressing barriers to and facilitators of PrEP uptake and adherence with a multi-level approach, incorporating personal, social, health systems and policy level factors. Though participants reported primarily on PrEP adherence as it related to their personal and social spheres, their views and experiences stressed the importance of systemic and policy interventions to promote the safe and effective use of PrEP. The acquisition of robust evidence about PrEP safety during pregnancy is of critical importance for the development of clear clinical guidelines and strategies for their implementation toward further control of HIV in the general population and the eventual elimination of mother-to-child transmission of HIV.

## Data Availability

Data supporting the conclusions of this article can be retrieved following the data sharing policy at the MRC/UVRI & LSHTM Uganda Research Unit. This policy can be assessed through the following link: https://www.mrcuganda.org/publications/data-sharing-policy.

## ACKNOWLEDGEMENTS

We are appreciative for the support of the community members in Kampala who generously gave their time to talk to us and we thank MRC/ UVRI and LSHTM Uganda Research Unit. We are particularly grateful to the POPPi team members for their work in keeping the projects alive through the COVID-19 pandemic. Thank you.

## ETHICAL APPROVALS

In Uganda approval was given by the Uganda Virus Research Institute Research Ethics Committee (GC/127/19/02/564), the Uganda National Council for Science and Technology (SS4479). At UCSF (IRB approval #322235).

## Notes

### Competing Interest Statement

The authors have declared no competing interest.

### Clinical Trial

NCT04030520

### Funding Statement

This work was funded by the National Institutes of Mental Health R34MH114523-01

### Author Declarations

In Uganda approval was given by the Uganda Virus Research Institute Research Ethics Committee (GC/127/19/02/564), the Uganda National Council for Science and Technology (approval number: SS4479). In the United States, approval was grated by the University of California, San Francisco (IRB approval #322235).

